# When Screening is a Death Sentence: A qualitative study of fear and delay of cervical pre-cancer screening in Kilombero District, Tanzania

**DOI:** 10.64898/2026.08.26.26361100

**Authors:** Margareth Somba, Mari Dumbaugh, Grace Mhalu, Sonja Merten, Sally Mtenga

## Abstract

**Background:** In Tanzania, over 10,000 women aged 15-44 are diagnosed with cervical cancer annually, and more than 6525 die. The majority are diagnosed at an advanced stage, which contributes to delays in treatment enrollment and illness complications. Multiple factors can affect women’s willingness and ability to access cervical pre-cancer screening.

**Purpose:** To explore the barriers and facilitators to attending cervical pre-cancer screening among women living in a resource-constrained area of Southern Tanzania

**Methods:** A qualitative study of 17 focus group discussions and 12 in-depth interviews was conducted from December 2023 to July 2024. Data collection took place in the community and health centers of a town and the surrounding rural areas in Kilombero district. The study used purposive sampling to recruit 112 women and 23 men aged 18-50+ years.

**Results:** Fear of death emerged as a central theme in the data, acting as both a barrier to and a facilitator of women’s screening behavior. Women linked death with the screening procedure, receiving results and undergoing treatment. Participants mistrusted the speculum itself and linked it to pain, vaginal infection and infertility. Conversely, fear of physical and social death motivated other women to attend the screening to learn about their health and prevent the consequences of a positive diagnosis. Public trust in the healthcare system and the role of health information sources were also identified as influencing women’s decisions to undergo or not undergo screening.

**Conclusion:** Our findings showed that cervical cancer screening uptake is influenced by fear of death and other co-factors. To improve early cervical cancer screening, programs and policy interventions are needed to raise awareness of the disease while also addressing women’s specific concerns. Also, strengthening structural dimensions such as the availability of the healthcare workforce, healthcare facilities and ensuring equitable service availability are essential to reducing cervical cancer complications and avoidable mortality.

## Introduction

Cervical cancer (CC) is the leading cause of cancer-related death among women worldwide. In 2020, there was an estimated 604,127 cases and 341,831 deaths globally, with approximately 90% of deaths occurring in low-and middle-income countries (LMICs) (1, 2). In 2022, 23.3% of new CC cases and 23% of global CC deaths were recorded in Sub-Saharan Africa alone (3, 4). Tanzania had the highest CC incidence and mortality rates among East African countries (5–8). By 2025, the incidence rate was projected to rise to 12,416 cases and 9,923 deaths per 100,000 women (9). The high mortality seen in Tanzania is due to ineffective control measures and can only be prevented through early screening and treatment of pre- cancerous lesions (10).

The primary risk factor for CC is the human papillomavirus (HPV), with 70% of all CC cases caused by HPV types 16 and 18 (11). Therefore, primary prevention of CC is achieved through the HPV vaccine, which is a safe and effective measure. Secondary prevention includes early screening and treatment of pre-cancerous lesions to prevent the development of invasive CC (12). Although screening identifies cases of invasive cancer, its primary purpose is prevention: Pre-cancerous lesions can be treated easily and at low cost, thereby preventing CC (13). For this reason, this paper refers to the intervention as cervical pre-cancer (CpC) screening.

To lower the burden of CC, the World Health Organization (WHO) has established the 90-70-90 strategy. This strategy ensures that by 2030: 90% of girls receive the HPV vaccine by age 15; 70% of women are screened, and 90% of women with pre-cancerous lesions receive treatment (14). Also, Sustainable Development Goal (SDG) Target 3.4 aims to reduce premature mortality from non-communicable diseases (NCDs) by one-third by 2030(8). To achieve these goals, the Tanzanian government, through the Ministry of Health, introduced the HPV vaccination program in 2014 for schoolgirls aged 9–14 years at no cost. To date, 79% of eligible girls have received a single dose, and 60% have received a second dose. Despite the success of the HPV vaccination campaign, CpC screening remains important because HPV lowers the risk of CC but doesn’t eliminate it (4).

While the WHO’s global target is to eliminate CC by 2030 through vaccination and early CpC screening, evidence shows that a small proportion of women in sub-Saharan Africa have been screened for CpC. The screening coverage ranges from 2.0% to 20.2% in urban areas and from 0.4% to 14.0% in rural areas, deterring efforts to control CC (15). In Tanzania, despite the expansion of screening programs, improved community awareness campaigns, and free screening services, the uptake of CpC screening remains low, at only 11% of women aged 30-50 (16–19). There is evidence of systemic and social barriers to CpC screening, including lack of awareness, limited infrastructure and resources, stigma and shame (20). We had similar findings but also fear of death emerged throughout the participants’ narratives. Existing literature has identified this theme but has not explored it in detail(20) To address this important gap in the literature, we explored fear of death as a barrier and facilitator to CpC screening.

## Methods

### Study design and setting

We conducted a qualitative study using in-depth interviews (IDIs), focus group discussions (FGDs) and observation to explore barriers and facilitators that influence the uptake of CpC screening. The study was conducted in Kilombero district, Morogoro region, southeastern Tanzania across three study sites: a town (urban) and two rural villages, one 22 km and the other 56 km away from the town. The population of the town could easily access essential healthcare, including CpC screening and treatment services, compared to the populations in the surrounding rural areas (21). The rural communities depended on CpC community outreach programs delivered in the local dispensaries.

### CpC and CC services in the study area

CpC screening services in the town were offered at a government secondary health facility and two private tertiary hospitals. The level of cervical cancer services among the three facilities varied considerably, with the government facility offering screening using Visual Inspection with Acetic Acid (VIA) and cryotherapy treatment. The private facilities offered a broader range of services, including CpC screening with VIA, assisted self-sampling, biopsy and treatment through cryotherapy and surgery. One of the private facilities provided more advanced care, including chemotherapy and radiation, and received referrals from the two other facilities. Patients with severe cases were referred to the National Tertiary Hospital in Dar es Salaam, about 416 km away, for advanced care.

The government health facility conducted facility-based CpC screening twice a week, while the private facilities conducted it five days a week. Additionally, the government facility and one of the private hospitals collaborated to conduct community outreach programs for CpC screening, based on the availability of funds. The community outreach program is a collaborative effort by the public and private health departments in town, in which nurses and doctors travel to rural communities to provide CpC screening. This program saved women time and travel costs to the specialized hospitals in town. Women received CC education, free screening, and referrals for cases that required further diagnostic and treatment services. At the time of data collection, these facilities received financial support from international organizations to run CpC screening activities. However, the community outreach program for CpC screening services is currently unavailable due to foreign aid cuts.

In contrast, at both village sites, local dispensaries lacked facility-based CpC screening services. No outreach activities were offered in the village closer to the town; the farther village received periodic community outreach programs supported by the development partners as well as during government celebrations such as the Uhuru Torch or AIDS Day that took place once a year. Women with dysplasia, or CpC, at these events were referred to the private generalist hospital in town for further diagnosis and treatment.

### Study population and sampling procedure

This study involved 122 women and 23 men aged 18 to 75. Purposive sampling was used to identify participants 18 years and older with a variety of experiences with CpC screening and cervical cancer. Women for IDIs were recruited from the general community within the study area and included both screened and unscreened individuals. Three groups of women were recruited for FGDs: (i) women from the general population not screened, recruited in their community; (ii) women who attended CpC screening without any identified lesions, recruited from CC screening records; (iii) women living with HIV, not screened for CC, recruited at ART clinics. Men were recruited from the general community in each study site. Participants’ selection criteria included being married and aged between 20 and 49 years.

The first author (MS) worked with community health workers (CHWs) to recruit participants from each site, as they are trusted and can address fears and mistrust. During recruitment, CHWs informed participants about the study objectives and provided those interested with the date, time, and venue for their scheduled IDIs and FGD sessions. MS, a Tanzanian PhD student, maintained daily contact with the CHWs at each site to arrange activities and ensure adherence to eligibility criteria.

### Data collection

Prior to formal data collection, GM (PhD, qualitative researcher and project coordinator) and MS conducted formative qualitative data collection comprising three FGDs, one per site, to help finalize the research tools. The research team modified questions based on the research objectives and the study participants’ comments and the topics were explored during data collection.

Data collection for the main study took place from December 2023 to January 2024. Before beginning formative and main study data collection, informed consent was obtained from all participants. Each participant signed two copies of consent forms, keeping one copy for their record. Those who could not read or write used a fingerprint in place of a signature. Data was collected in the local language, Swahili, in private rooms in government schools and offices. These places were easily accessible and preferred by participants. MS trained in qualitative methods and conducted IDIs and FGDs with the help of a Tanzanian researcher with a bachelor’s degree in Sociology. All data were audio-recorded with participants’ permission. The IDIs lasted for 45–90 minutes, while FGDs lasted for 90 to 120 minutes. A semi-structured interview guide supported the exploration of the participants’ understanding of cervical cancer, their health-seeking behavior, barriers and facilitators to screening, and consequences of a positive screening result. The research team conducted 12 in-depth interviews and 17 FGDs until data saturation was reached.

Additionally, six non-participant observations were conducted around health facilities. These observation sessions took place during CpC screening in a private room designed for this purpose. After each session, MS took notes on the general screening environment, screening procedure, post-screening results, feedback and interpersonal clinic communication between the provider and the client.

In August 2026, we conducted dissemination meetings with study participants across urban and rural areas, as well as regional and district government representatives to share and discuss the findings.

### Data analysis

All audio files were labeled with non-identifiable participant IDs and stored on a secure local server. Trained researchers translated and transcribed the Swahili audio-recorded data verbatim into English for analysis. The project coordinator (GM) ensured data security during transcription and translation. MS reviewed the transcripts for coherence and identified areas for correction. All transcripts were anonymized and saved to local and Swiss-based secure servers, accessible only to limited members of the study team with a password.

Using an inductive approach, MS and MD (PhD and qualitative expert) read through transcripts for familiarization and identified preliminary codes. MS then inductively developed themes by coding the first group of three interviews, formulating an initial code book and sharing it with qualitative team members who double-coded the first group of interviews (Principal Investigators and PhDs, SMt, SMe; GM and MD). The qualitative team met to discuss, refine and finalize the codebook, resolving differing opinions via consensus. The MAXQDA (version 24.6.0) software program was used for data management and to organize analysis.

### Ethical approval

Ethical approval was obtained from the institutional review boards of Swiss TPH and Ifakara Health Institute, Tanzania, the “Ethikkommission Nordwest-und Zentralschweiz” (EKNZ) in Switzerland, and the National Institute for Medical Research (NIMR) in Tanzania.

## Results

### Characteristics of study participants

A total of 112 women and 23 men with ages ranging from 18 to 75 years participated in the study. Most of the participants (63%) had primary education, and 51.9% were married. About 42.2% of the participants reported belonging to a community or mutual support group where they occasionally shared information about health issues, including CC. (Table 1).

**Table 1:** Demographic information of study participants (N=135)

| **Characteristics** | **Interview** | **FGD** | **All Participants** |
| --- | --- | --- | --- |
|  | **Total (N=12) n (%)** | **Total (N=123) n (%)** |  |
| **Age** |  |  |  |
| 18-29 | 7(58.3) | 42(34.1) | 49(39.30) |
| 30-49 | 3(25.0) | 39(31.7) | 42(31.11) |
| 50+ | 2(16.7) | 42(34.1) | 44(32.59) |
| **Sex** |  |  |  |
| Female | 12(100) | 100(81.3) | 112(82.96) |
| Male | 0(0) | 23(18.7) | 23(17.04) |
| **Marital status** |  |  |  |
| Single | 4(33.3) | 31(25.2) | 35(25.93) |
| Married | 6(50.0) | 64(52.0) | 70(51.85) |
| Divorced | 1(8.3) | 15(12.2) | 16(11.85) |
| Widow | 1(8.3) | 13(10.6) | 14(10.37) |
| **Education** |  |  |  |
| No education | 0(0) | 11(8.9) | 11(8.15) |
| Primary | 6(50) | 79(64.2) | 85(62.96) |
| Secondary | 5(41.7) | 28(22.8) | 33(24.44) |
| Tertiary | 1(8.3) | 5(4.1) | 6(4.44) |
| **Occupation** |  |  |  |
| Skilled | 3(25) | 17(13.8) | 20(14.81) |
| Unskilled | 9(75) | 97(78.9) | 106(78.52) |
| None | 0(0) | 9(7.3) | 9(6.67) |
| **Support group** |  |  |  |
| Yes | 7(58.3) | 50(40.7) | 57(42.22) |
| No | 5(41.7) | 73(59.3) | 78(57.78) |
| **Ever screened for CC** | 2(16.7) | 39(39.0) | 41(36.61) |

Through thematic analysis, we developed three main themes linking fear of death to CpC screening behavior: the lack of clarity of the curability of CC, lack of trust in the quality of health services and social death and the threat to personhood. These factors acted as both barriers and motivators for screening uptake for different women. While our findings triangulated the existing literature which highlights the influence of structural factors such as lack of knowledge about CC, limited infrastructure/resources, and long distance to screening centers on access to care (22, 23), this paper specifically focuses on the fear of death and women’s decision to access CpC screening. For some women, the fear of death was a barrier to accessing CpC screening, especially if they believed CC was incurable. Conversely, other women, fearing death by CC but perceiving it as a treatable disease, were driven to access screening and preventative services to avoid an advanced CC diagnosis.

### The lack of clarity of the curability of C(p)C

Fieldwork revealed that CpC was not distinguished from CC. For example, screening for CpC was referred to in the health system and the community as ‘CC screening,’ and most women did not know that CpC treatment was available and free in select public health facilities in the region. Some women believed that ‘CC’ was incurable regardless of the stage in which it was diagnosed. In this context, our results illustrated that the perception of curability is socially constructed, grounded in women’s perceptions, personal experiences or observations of others. This view was driven by community narratives that women receiving CC treatment inevitably died during the treatment process or soon after; the treatment only provided relief and not a cure. An older woman farmer from the rural areas with a primary education shared her perspective about the effectiveness of CC treatment:

> Chemotherapy does not cure CC. It provides relief for a short time, and then the patient dies. A significant percentage of patients who receive chemotherapy die. (Woman, never screened)

This belief was compounded by the fact that many women sought screening or medical care only once symptomatic, by which time the disease had progressed to stage 3 or 4. Some delayed access to screening due to long distances and financial constraints for transportation and additional tests. Additionally, during dissemination meetings, participants shared that live radio broadcasts discussing CC invited listeners to call in with questions or comments. Some listeners shared sad stories or testimonials of their difficult time as caregivers for CC patients, or similar stories they heard about patients in their community. Therefore, community narratives emphasized the poor treatment outcomes and then death for patients in stages 3 and 4 of CC more than successful treatment of pre-cancer and cancer, reinforcing the perception that CC is incurable.

Some women went further and questioned the utility of screening if they could not afford the treatment for precancerous conditions and the expensive follow-up for advanced cancer. For example, a participant from an urban area explicitly shared her perspective about the uncertainty of treatment, although she had undergone screening:

> We often see people on the street who have CC, but they don’t get anywhere despite the treatment; survivorship is minimal, and they eventually die. That is the biggest thing that many of us in the street are afraid of. “Aah, this one has CC; it will take her slowly at the end, she will just die”. (Woman, screened)

Another participant, who perceived the disease as incurable and fatal, was concerned about the efficacy of screening, viewing the process as a waste of money and time. She argued that prioritizing screening for a fatal disease over household chores was unreasonable, especially while she remained asymptomatic and healthy. Residing in a rural setting, where access to screening required money and time to travel to town, she reported that:

> As a human being, I feared dying prematurely if diagnosed with CC. I didn’t know if I would survive because the disease is dangerous and many people have died from it. Even getting screened at a large hospital is just a waste of money. If I am healthy, why would I spend money on screening? I feel like I am delaying continuing with my activities. (Woman, never screened)

The fear of an uncertain future was not confined to individual women, but a collective anxiety shared by families and communities. Traditionally, women play a vital role in providing for their families and are expected to farm, care for children, and handle other household chores. A positive CC diagnosis represented more than a medical condition; it signified being out of production, directly jeopardizing the family’s livelihood. Subsequently, for some women, the decision to opt out of screening was an attempt to stay productive and maintain family stability. A participant from a rural community, reported her hesitancy for screening was rooted not only in the fear of her own death but also the uncertain future for their children.

> Before going for CC screening, I was in constant fear; I didn’t know what would happen to my children. Who would support them? Would I survive and be able to care for my children? My mother advised me to get screened, but it took me two months to decide. My fear was about the future of my children if I were diagnosed with CC, would it be cured, or would it be like HIV? (Woman, screened)

In contrast, some participants believed that CC was curable if diagnosed and treated in its early stages, acknowledging that the survival rate was minimal if a woman delayed and sought care when the disease had developed into invasive cancer. These participants emphasized hospital treatment over other options like traditional medicine, prayers or self-medication for better outcomes. Many participants reported their initial visit was at the lower-level health facilities (dispensary) in response to symptoms such as vaginal bleeding, pelvic pain, or discharge that resulted in misdiagnosis and treatment for urinary tract infections. It was after several follow-ups that these women were referred to and accessed the appropriate facilities for diagnosing and treating precancerous conditions.

> …If you are diagnosed with a disease and get treatment early, you can be healed completely. But once you opt for traditional medicine, the disease will progress to an advanced stage, making it difficult to treat. (Woman, screened)

Participants also noted that witnessing positive treatment outcomes could serve as a powerful motivator for women who had not screened to do so, as one older woman participant shared her perspective:

> If a woman is diagnosed with the disease, is treated and recovers, the rest of us here will go for screening because it proves the disease is treatable when screening is done early. (Woman, screened)

### Varying levels of trust in the quality of health services

Another major theme we developed during analysis was participants’ consistent lack of trust in the health system’s ability to deliver quality services, ranging from a negative view to total distrust. The mistrust was driven by experiences or perceptions of poor clinical procedures that were reported as painful or perceived to carry a risk of infection. These negative experiences triggered fear, with many participants equating the screening process with the risk of death.

### Perceived painful screening

Fear of the screening procedure was cited by many women, particularly the use of the speculum. Women who had not undergone screening perceived the insertion of the speculum as not only painful but also causing long-term physical harm, such as cervical enlargement and infertility. These fears were validated by community narratives shared between women, which described the clinical procedure as something to be avoided. For most women, the decision not to screen was solidified by hearing about others’ negative experiences. A woman from a rural area, who had never been screened, reported how the negative encounter at the health facility changed her decision to screen:

> It was scary; I ran away from the hospital…. I didn’t go to the screening room. I found a woman coming out of the screening room, complaining, ‘If I had known, I wouldn’t have come! She cautioned me! Just leave; this is not the place to come. They will insert a big iron metal (speculum) into your vagina. I gave up and said I’d rather die than go for a screening. (Woman, never screened)

This experience illustrates an observed pattern in which negative stories shared within the community become a powerful tool for instilling fear and causing women to opt out of screening.

Participants also linked the use of a speculum with infertility. They believed that during the screening procedure, the providers pushed on the uterus, damaging the cervix for conception. A woman from an urban area who had never been screened shared the community’s perspective on screening:

> When women go for screening, the provider inserts a speculum that looks like a ‘duck’s mouth’, which causes a lot of pain. A woman will not be able to conceive after the procedure because the provider pushed the uterus with the speculum and blocked it. (Woman, never screened)

This fear is embedded in a cultural context where parenthood is the most desired and valued goal for married couples. Moreover, the community expects couples to have a child in the first year or two years of marriage; if that does not happen, the woman is often blamed. In these settings, children are highly valued as they ensure the continuity of the family name (patrilineal), stabilize marriage and are believed to take care of parents when they grow old. Due to society’s definition of infertility, women who do not have children are victims of community rejection and divorce, although infertility can affect both men and women. Therefore, avoiding the speculum, regardless of who was performing the procedure, was an important strategy for women to protect their social personhood.

### Perceived risk of infection

Mistrust was also related to the speculum used during community outreach programs. It was not uncommon for 100-200 women to show up for community screening days. On such occasions, some participants believed that the providers used a single speculum for all women without sterilizing the equipment between patients; as a result, some left the screening area or avoided screening altogether for fear of contracting vaginal infections.

> I remember when I went for screening, the provider used the speculum that I heard other women were screened with. I lived with the fear of vaginal infection, but I have not experienced vaginosis to date. (Woman, screened)

Others, such as this woman who was never screened, were discouraged by negative community narratives and a perception that the screening procedure itself could cause CC:

> When I went to the facility for screening, someone discouraged me, saying the procedure could cause CC. (Woman, never screened)

According to the community CpC outreach team, each speculum was used per procedure and then sterilized before being reused. The team reported carrying 100 speculums, and once 50 had been used, they placed them in a portable autoclave machine for sterilization while continuing with the screening. After 30 minutes of sterilization, the speculums were reused, and the sterilization procedure was repeated depending on the client’s flow. Community outreach programs were conducted in health facilities with reliable electricity to facilitate the screening procedure. During the site observation conducted as part of this research, sterilization of speculums was not necessary because only a small number of women attended the screening and there were enough previously sterilized speculums to be used.

While hesitation to attend screening was widespread, we did encounter women who trusted the quality of health service provided and found screening beneficial, not a threat to their health, as exemplified below by a woman from rural who attended screening:

> I would say we are very grateful because in the past, they came for testing, but CC was not on the list [of screening services offered]. We are thankful that our medical experts are coming to examine women in the countryside and provide us with this service because it saves us the cost of going to [town]. (Woman, screened)

Notably, few women who had experienced both CC screening methods, self-sampling and VIA, reported that self-sampling was more convenient, easier to use, and more comfortable (less painful) than other clinician-assisted sampling methods, as described by this woman with both screening experiences:

> Aaah! Self-testing is painless; you insert the device yourself, slowly, until it reaches the area the provider directed. Conversely, when the doctor runs the VIA procedure, she inserts the device directly into the cervix without considering the pain you may experience. I think it’s best to examine yourself. (Woman, screened)

### Providers’ communication

The majority of participants who underwent screening reported that positive interactions with healthcare providers when seeking care facilitated trust in health services. For example, the provider’s effective communication was observed during the screening services at a public facility in town. The provider used verbal techniques to manage patients’ anxiety, such as calm language, saying ‘sorry’ or ‘relax’ when inserting the speculum or ‘I am now inserting the speculum’ to prepare the woman mentally for the procedure. The rapport calmed women and dispelled fear, motivating them to access screening, as narrated by a woman from town:

> Eeeh, providers were skillful; they showed us a place to sit and wait for the doctor. There is nothing better for a patient than good treatment. I felt unique because I went there without any sickness, and the doctor encouraged me to get screened, even though it’s my health. (Woman, Screened)

Patient satisfaction during screening site observations was evident as women expressed gratitude after the screening procedure. Participants from both sites reported on a supportive environment, including clear instructions on where to sit, recommended follow-up visits, and encouragement

### Fear, social death and the decision to access CpC screening

Fear was a central, multifaceted theme in women’s CpC decision-making. Participants specifically referenced fear of death, which could act as either an obstacle or a motivating factor for women’s screening attendance. For some women, fear prevented them from accessing screening. These women explicitly expressed fear of the word cancer, the CpC screening process, of knowing the results, and of undergoing treatment.

> The name cancer is scary to people; they associate it with premature death; a person sees that it is the end of their life. (Woman, never screened)

In discussing their screening choices, women linked their fears of the screening process, receiving positive results, and undergoing treatment to their fear of death. Consequently, participants’ decisions to undergo CpC screening were shaped by their conceptualization of death, which was defined in three dimensions: physiological, social and spiritual. Some Physiological, women simply referred when the vital organs stop working, as explained below:

> Death is when a person’s breathing stops and they cannot do anything. (Woman, never screened)

Other women discussed *spiritual* death, meaning the death of the soul. It was also associated with life after death

> Death is scary. You die, the soul leaves the body, and it doesn’t come back. You don’t know where the soul goes (laughter) I’m scared! If you think of what the scriptures say about death, you will be afraid. The Bible says: When you die, two angels of good deeds and evil come to your grave and judge you accordingly. (Woman, never screened)

In this sense, death was conceived as something inevitable, the final celebration alongside birth and marriage. Others referred to the biblical teachings about disappearing from the world without knowing where you are going.

While spiritual and physical death were inevitable and linked to the end of someone’s life, another dimension, *social death,* emerged as a powerful threat that only befell certain people. *Social death* was described as a process that begins much earlier than physical death and was never conceived in a celebratory or positive light. *Social death* has been described in the literature as occurring when a family or community withdraws from, isolates or rejects a vulnerable person and considers them dead while they were still alive, a concept that has been linked to patient experiences across diseases, but especially those experiencing chronic disease (24).

Women in our study diagnosed with CC described experiencing social death, a process through which they prematurely lost their identity and position in the community. As the disease progressed, the patient was seen as a burden to the family and the community because she could no longer fulfil essential roles such as farming, household management or marital duties. At this stage, close people such as family, relatives or partners distanced themselves and stopped supporting her out of fear of being infected or to avoid financial or emotional burden. One participant shared a negative experience of her friend, who experienced isolation and abandonment from the family:

> I know a woman with CC whose husband ran away when she was about to start the treatment. Her relatives came and looked after her for a while and then stopped the care, no one came to comfort or support her until she died. (Woman, never screened)

This negative experience slowly killed the patient emotionally and psychologically before her physical death. She lost a sense of attachment and belonging in her family.

Observations of women who experienced stigma due to their CC condition were common. Similar to social death, stigma manifested as social exclusion, avoidance, and care-taking fatigue from the family and close relatives. For example, one participant shared a negative experience of her friend, who was living with a chronic disease and was later diagnosed with CC. She complained about experiencing isolation from her family after her CC diagnosis.

> I remember talking to my friend who was diagnosed with chronic diseases. She was down, crying, and had lost hope. She said she was going to die, and her children were tired of taking care of her. I told her not to give up and to trust in God. I guess she gave up psychologically when she saw that they had separated [her] serving dishes [from the family’s]; they thought they would be infected. (Woman, never screened)

For some women, a positive CC diagnosis led to abandonment by their spouses or divorce once they, as wives, were perceived as economically unproductive or sexually incapable. Marriage is considered important for intimacy, procreation, and economic and social support. In some communities, married women gain identity and are treated with more respect than unmarried women. For example, women from the study sites reported that their primary activity is farming maize and rice. During the agricultural season, they are expected to plant, weed, spray pesticides and harvest. When the husband went to the farm, the wife was expected to work alongside him. In other families, it is only the woman who works in the field while the husband engages in other economic activities, such as running a small business or driving a motorcycle. In such instances, women must work hard to harvest enough food to feed the family and sell the surplus to cover other basic needs. As elaborated by a woman with four children, who made a living by herself after her marriage ended:

> We rely on farming for survival; if I am diagnosed with CC, I would be stressed out thinking about the disease and farming. I would be concerned about who will take care of my children and the farm if my condition gets worse. Nobody would take charge, not even relatives. Sometimes you feel it’s best not to screen for peace of mind. (Woman, never screened)

Moreover, women are expected to look after their reproductive health for procreation and the welfare of their husbands and children. Therefore, some women felt it was best not to screen to avoid devastating consequences of a positive diagnosis, such as worrying about who would look after them during sickness, who would take care of the field and provide for their children, and they also wanted to preserve their marriages.

Women also shared that they feared a positive CC diagnosis because of the shame they would feel for failing to “protect” their cervixes and fulfill their expected roles within their families and community. A middle-aged woman from a rural area shared her perspective:

> Women are expected to maintain good hygiene, practice safe sex, eat healthy and get regular checkups to protect their cervix… It’s challenging for young women to handle the consequences of a positive result. If I were single and diagnosed with CC, there is a chance that I wouldn’t get married. People would consider me a dead person regardless of the disease stage, feeling that, due to my condition, I cannot reproduce and that treatment may shorten my life. (Woman, never screened)

This notion is rooted in societal expectations regarding women’s role in protecting their cervix for reproduction. Participants noted that the community views CC and other reproductive health issues primarily as women’s problems; she is responsible for protecting herself against any unexpected health problem.

For some women, witnessing such experiences instilled fear in them and acted as a barrier to CpC screening. Fear of experiencing social death drove these women away from CpC screening to preserve their social personhood. Participants stated that maintaining their current social relationships – their personhood, marriage and place in the community – was more important than the risk of being diagnosed with CC and losing their family bond, mental stability, and even marriage.

For other women, however, fear of social death was a motivator to go for CpC screening rather than a source of fear and anxiety. After witnessing their relatives or friends suffering due to stigma, lack of support, divorce and ultimately death, observation of negative experiences from loved ones was an inspiration to go for screening to be aware of their health, protect their cervix and avoid negative consequences of a positive diagnosis. They emphasized that early screening allows immediate treatment of observed symptoms, thereby preventing further infections and avoiding the social and economic burden of late-stage diagnosis. For example, this woman shared that she was motivated to go for CC screening after witnessing what her aunt had experienced because of her CC condition:

> My sister was abandoned by her husband. Her relatives took care of her, but they grew tired and left my aunt to care for her young children. She received radiotherapy for 6 months, but her condition did not improve, so the doctor discharged her. Sadly, she died a week later. She lost hope following the stigma from the family. (Woman, Screened)

Another participant shared a decision to screen due to her aunt’s struggle with physical pain and social rejection due to invasive cancer. The aunt was enrolled in chemotherapy, but her condition did not improve; when she was discharged from the hospital, she opted for traditional medicine. Unfortunately, she passed away during the process of receiving care. Witnessing her aunt’s negative experience changed the participant’s perspective on the disease, prompting her to seek screening:

> … My aunt died of CC, and before dying, I heard her complaining about the physical pain and social exclusion she experienced. I was convinced to go because of such an experience. It (CC) is a dangerous disease. (Woman, screened)

## DISCUSSION

CC is a critical public health problem, with a high number of deaths concentrated in LMICs. While various factors have been cited as barriers to early CpC screening (16, 25, 26) this study found that fear of death was one of the main narratives in women’s accounts of why they would not want to use CpC screening. This fear was multifaceted and was informed by many factors that shape women’s healthcare-seeking, from structural to social-interactional and emotional aspects. Specifically, the screening uptake was influenced by a lack of clarity about the curability of CC, mistrust in the quality of health services, high cost of services, and fear of social death and the threat to personhood.

Participants in this study expressed hesitation to go for screening due to fear of the disease, equating the word ‘cancer’ to death. Women also feared the screening procedure itself, receiving test results, and treatment that was often perceived as categorically unaffordable. These findings are in line with previous studies from Tanzania and China, where women delayed screening due to fear of positive test results and treatment costs, gearing their preference towards traditional medicine or prayers (27, 28). Our findings expand the understanding of women’s fear of CC to include fear of social death if diagnosed with CC. Women with a CC diagnosis experienced social death from close social networks driven by the fears of treatment costs, financial burden, emotional strain and care fatigue at the household level, and misinformation about disease transmission. Consequently, these women were isolated, abandoned and treated as dead while they were alive, an experience that has been documented with other chronic diseases(24).

CC’s perceived threat to social position and personhood motivated some women to screen and others to opt out. Women in Tanzania are expected to contribute to household economics, including housework and childcare and, in rural areas, fieldwork from planting to harvest. This is also the case with other African settings, where marriage, children and the economic role of women are highly valued (29, 30) This is also the case with other African settings, where marriage, children and the economic role of women are highly valued. Some women, however, did not undergo screening due to the fear of negative consequences of a positive diagnosis, such as stigma, abandonment or divorce (26, 31). However, our study also showed that other women opted for screening to learn about their health. Negative results assured women of their ability to perform their social roles, or, in the case of a positive CpC result, early treatment would help them remain socially and economically functional. A study in Kenya had similar results; women screened to learn about their health and avoid living with the fear of having CC (32).

Participants in this study were also hesitant about CpC screening because a diagnosis of dysplasia, or pre-cancer, was perceived to be a diagnosis of cancer itself. In our fieldwork, CpC screening programs were commonly referred to as ‘CC screening,’ and cancer was thought to be incurable, regardless of the stage. This triangulates findings from a study in Kenya where women also perceived CC to be incurable (32). Indeed, if women delayed screening until the appearance of physical symptoms, access to treatment and treatment outcomes remained poor and could result in death. This made CC-related deaths more visible in our study community than healthy recoveries (33). Therefore, if women are well informed about the role of CpC screening in preventing CC, if screening is easily accessible in nearby facilities, and if treatment for CpC and CC is accessible and affordable, women might be more ready to consider screening. Community outreach programs were considered an effective way to screen many women, and participants appreciated saving time and transportation costs to go to facilities in town (34). However, foreign aid cuts have significantly affected the outreach activities. Therefore, government prioritization of cervical cancer screening and allocating funds to support outreach screening services will be crucial to make screening available to all women. In addition, achieving universal health coverage with a package that includes pre-cancer and cancer treatment would alleviate financial constraints that contribute to women’s fear of screening, thereby increasing the perceived utility of screening.

Women also reported avoiding CpC screening out of fear of pain during the procedure or long-term physical damage such as enlarging the vagina and abrasion that would cause CC, or infertility (29, 32, 35). Similarly, studies in Kenya and Guatemala reported that some women would not undergo screening due to the fear of pain or discomfort during the screening (32, 36). Beliefs that insertion of a speculum damaged the cervix or pushed the uterus, deterring reproduction and posing a threat to women’s societal role, were also observed in other settings (25, 37). Cross-sectional survey data of women who went for screening in the same study area show that most women had positive screening experiences (Kipo, forthcoming). However, our data raises the question of how some women, albeit the minority, are experiencing screening services. For example, rushing examinations because of high patient volume or inserting a speculum without talking to the patient through the procedure, gaining her consent, or using lubrication could cause pain or injuries. Women with vaginal or cervical inflammation could also find pelvic exams painful. Some women may have been mistreated or not treated with respect in the health system, so they are scared now to go back.

Our findings highlighted the value of interpersonal clinical communication in facilitating positive clinical experiences. We observed that empathetic verbal communication, such as using calm language and expressing reassurance of the screening procedure, reduced fear in women undergoing screening. These findings suggest the need for comprehensive education that addresses context-specific fear and beliefs about CC screening. For example, information campaigns could also make the screening process more transparent for women, walking them through the examination procedure and reassuring them that the procedure does not have long-term health risks. Specific training for health providers in trauma-informed pelvic care could also improve the patient experience by ensuring informed, safe, comfortable and empowering patient-provider interactions and exams (38).

In addition, individuals, as social entities, are influenced by shared experiences and cultural norms, either positively or negatively. Notably, this study and others (39, 40) found that hearing or witnessing negative consequences of CC disclosure instilled fear in unscreened women, discouraging them from going for screening for fear of experiencing the same negativity. Engaging women’s social networks to diffuse positive screening experiences throughout the community could be an effective strategy. These findings also suggest the need for media campaigns to increase knowledge of the risk factors and causes of CC, the benefits of early, pre-symptomatic screening and later stage CC symptoms. To leverage the power of social networks and positive experiences to influence health decisions, community education campaigns could feature testimonies from women who experienced CpC screening and CC survivors, distinguishing CpC from CC, explaining the screening process is not painful and that CpC and CC are curable if detected at an earlier stage. Such stories could reduce fears surrounding screening and address any beliefs that contribute to low screening uptake.

## Strengths and limitations of this study

This study explored fear of death and co-factors influencing the uptake of CpC screening, which are not widely addressed in the literature, by applying a health social science lens to this challenge. We interviewed a diverse group of key stakeholders while centering women’s experiences, and incorporated several data collection methods to triangulate findings, including individual interviews, focus groups and site observations. Data collection was completed in Swahili by a Tanzanian researcher familiar with the context, and positive rapport with participants was prioritized to reduce social desirability bias. One limitation of our study was the challenge of recruiting women living with HIV for the study, who are, therefore, under-represented in our participant group. And, as with all qualitative research, our findings are only relevant to our specific study context.

## Conclusion

The findings of this study demonstrate that the fear of death was a central theme that influenced CpC screening behavior. Community perceptions which do not distinguish between CpC and CC, and equating CC with death, intensified the association between a C(p)C diagnosis and social death among women. To improve the uptake of CpC screening, community health education should engage with and respond to, rather than dismiss, local perceptions of fear: for example, public awareness can be raised through community meetings or mass media communications during which healthcare providers share key messages, the screening process is made more transparent and women treated for dysplasia and CC survivors share their experiences of screening, diagnosis and recovery.

## Data Availability

All data produced in the present study are available upon reasonable request to the authors

## Acknowledgements

We would like to thank all study participants for agreeing to participate in the study and share their experiences. Also, the health facilities and schools that offered space to conduct this research.

## Authors’ contributions

SMe developed the original study, obtained funding and revised the manuscript. SM contributed to the design of the study protocol and revised the manuscript. MS participated in data collection and writing of the manuscript. MD participated in study design and revised the manuscript. GM supervised data collection and revised the manuscript. All authors read and approved of the final manuscript.

## Availability of data and materials

The datasets used and/or analyzed during the current study are available from the corresponding author obtained upon request.

## Ethics approval and consent to participate

Ethical approval was obtained from the institutional review boards of Swiss TPH, Switzerland and Ifakara Health Institute, Tanzania. The “Ethikkommission Nordwest-und Zentralschweiz” (EKNZ) in Switzerland, and the National Institute for Medical Research (NIMR) in Tanzania. The ethical permission reference number NIMR/HQ/R.8a/Vol.IX/4408. All participants received information about the study and signed written informed consent to participate in the study.

## Consent for publication

Participants were notified of the future publication of results during the informed consent process.

## Author details

Society, Gender & Health Unit, Department of Epidemiology and Public Health, Swiss Tropical and Public Health Institute, Kreuzstrasse 2, 4123 Allschwil, Switzerland University of Basel, Peterspiatz 1, Postfach, 4001 Basel, Switzerland Ifakara Health Institute, P. O. Box 78373, Mikocheni B, Dar es Salaam, Tanzania.

## References

1. Singh D, Vignat J, Lorenzoni V, Eslahi M, Ginsburg O, Lauby-Secretan B, et al. Global estimates of incidence and mortality of cervical cancer in 2020: a baseline analysis of the WHO Global Cervical Cancer Elimination Initiative. Lancet Glob Health. 2023;11(2):e197–e206.

2. Sung H, Ferlay J, Siegel RL, Laversanne M, Soerjomataram I, Jemal A, et al. Global Cancer Statistics 2020: GLOBOCAN Estimates of Incidence and Mortality Worldwide for 36 Cancers in 185 Countries. CA Cancer J Clin. 2021;71(3):209–49.

3. Bray F, Parkin DM, Gnangnon F, Tshisimogo G, Peko J-F, Adoubi I, et al. Cancer in sub-Saharan Africa in 2020: a review of current estimates of the national burden, data gaps, and future needs. The Lancet Oncology. 2022;23(6):719–28.

4. WHO. African Region/ Health topics/Cervical cancer 2025 [Available from: https://www.afro.who.int/health-topics/cervical-cancer.

5. Cancer IIICoHa. Kenya Human Papillomavirus and Related Cancers, Fact Sheet 2023. 2023.

6. Cancer IIICoHa. Uganda Human Papillomavirus and Related Cancers, Fact Sheet 2023. 2023.

7. Cancer IIICoHa. Tanzania Human Papillomavirus and Related Cancers, Fact Sheet 2023. 2023.

8. WHO. Noncommunicable diseases: fact sheet on Sustainable Development Goals (SDGs): health targets 2017 [Available from: https://www.who.int/news-room/fact-sheets/detail/noncommunicable-diseases.

9. Henke A, Kluge U, Borde T, McHome B, Serventi F, Henke O. Tanzanian women s knowledge about Cervical Cancer and HPV and their prevalence of positive VIA cervical screening results. Data from a Prevention and Awareness Campaign in Northern Tanzania, 2017 - 2019. Glob Health Action. 2021;14(1):1852780.

10. Ngwa W, Addai BW, Adewole I, Ainsworth V, Alaro J, Alatise OI, et al. Cancer in sub-Saharan Africa: a Lancet Oncology Commission. Lancet Oncol. 2022;23(6):e251–e312.

11. CDC. Human Papillomavirus (HPV). Genital HPV Infections— Fact Sheet. 2022.

12. Wright K, Aiyedehin O, Akinyinka M, Ilozumba O. Cervical cancer: community perception and preventive practices in an urban neighborhood of Lagos (Nigeria). International Scholarly Research Notices. 2014;2014(1):950534.

13. WHO. Cervical cancer 2024 [Available from: https://www.who.int/news-room/fact-sheets/detail/cervical-cancer.

14. WHO. Global strategy to accelerate the elimination of cervical cancer as a public health problem: World Health Organization; 2020.

15. Yimer NB, Mohammed MA, Solomon K, Tadese M, Grutzmacher S, Meikena HK, et al. Cervical cancer screening uptake in Sub-Saharan Africa: a systematic review and meta-analysis. Public Health. 2021;195:105–11.

16. Mahiti GR, Adam J, Luoga P. Prevalence and determinants of cervical cancer screening among women aged 15–49 years in Tanzania: analysis of demographic and health survey 2022. Archives of Public Health. 2025;83(1):135.

17. Hall MT, Smith MA, Simms KT, Barnabas R, Murray JM, Canfell K. Elimination of cervical cancer in Tanzania: Modelled analysis of elimination in the context of endemic HIV infection and active HIV control. Int J Cancer. 2021;149(2):297–306.

18. Welfare MoHaS. National Cervical Cancer Prevention Control Strategic Plan (2011-2015). 2011.

19. WHO. Costing the National Response to Cervical Cancer: United Republic of Tanzania, 2020—2024. 2020.

20. Bateman LB, Blakemore S, Koneru A, Mtesigwa T, McCree R, Lisovicz NF, et al. Barriers and Facilitators to Cervical Cancer Screening, Diagnosis, Follow-Up Care and Treatment: Perspectives of Human Immunodeficiency Virus-Positive Women and Health Care Practitioners in Tanzania. Oncologist. 2019;24(1):69–75.

21. NBS. City Population Census 2022 [Available from: https://www.citypopulation.de/en/tanzania/coastal/admin/mlimba/105031161_idete/. .

22. Mtuya CC, Cadstedt J, Mattsson J, Manhica HA, Serventi F, Machange R, et al. “ Cervical Cancer—A Silent Disease in the Community”—A Qualitative Study on Awareness of Cervical Cancer in Tanzania. Sage Open Nursing. 2025;11:23779608251393079.

23. Mugassa AM, Frumence G. Factors influencing the uptake of cervical cancer screening services in Tanzania: A health system perspective from national and district levels. Nursing Open. 2020;7(1):345–54.

24. Ghane G, Shahsavari H, Zare Z, Ahmadnia S, Siavashi B. Social death in patients: Concept analysis with an evolutionary approach. SSM-Population Health. 2021;14:100795.

25. Kirubarajan A, Leung S, Li X, Yau M, Sobel M. Barriers and facilitators for cervical cancer screening among adolescents and young people: a systematic review. BMC women’s health. 2021;21(1):122.

26. Adedimeji A, Ajeh R, Pierz A, Nkeng R, Ndenkeh JJ, Fuhngwa N, et al. Challenges and opportunities associated with cervical cancer screening programs in a low income, high HIV prevalence context. BMC women’s Health. 2021;21(1):74.

27. Yang H, Li S-P, Chen Q, Morgan C. Barriers to cervical cancer screening among rural women in eastern China: a qualitative study. BMJ open. 2019;9(3):e026413.

28. Bateman LB, Blakemore S, Koneru A, Mtesigwa T, McCree R, Lisovicz NF, et al. Barriers and Facilitators to Cervical Cancer Screening, Diagnosis, Follow-Up Care and Treatment: Perspectives of Human Immunodeficiency Virus-Positive Women and Health Care Practitioners in Tanzania. The Oncologist. 2018;24(1):69–75.

29. Baloyi GT. Marriage and culture within the context of African indigenous societies: A need for African cultural hermeneutics. Studia Historiae Ecclesiasticae. 2022;48(1):1–12.

30. Oketch SY, Kwena Z, Choi Y, Adewumi K, Moghadassi M, Bukusi EA, et al. Perspectives of women participating in a cervical cancer screening campaign with community-based HPV self-sampling in rural western Kenya: a qualitative study. BMC Womens Health. 2019;19(1):75.

31. Morse RM, Brown J, Gage JC, Prieto BA, Jurczuk M, Matos A, et al. “Easy women get it”: pre-existing stigma associated with HPV and cervical cancer in a low-resource setting prior to implementation of an HPV screen-and-treat program. BMC Public Health. 2023;23(1):2396.

32. Buchanan Lunsford N, Ragan K, Lee Smith J, Saraiya M, Aketch M. Environmental and Psychosocial Barriers to and Benefits of Cervical Cancer Screening in Kenya. Oncologist. 2017;22(2):173–81.

33. Massobrio R, Bianco L, Campigotto B, Attianese D, Maisto E, Pascotto M, et al. New frontiers in locally advanced cervical cancer treatment. Journal of Clinical Medicine. 2024;13(15):4458.

34. Srinivas V, Herbst De Cortina S, Nishimura H, Krupp K, Jayakrishna P, Ravi K, et al. Community-based Mobile Cervical Cancer Screening Program in Rural India: Successes and Challenges for Implementation. Asian Pac J Cancer Prev. 2021;22(5):1393–400.

35. Boivin J, Carrier J, Zulu JM, Edwards D. A rapid scoping review of fear of infertility in Africa. Reprod Health. 2020;17(1):142.

36. Bevilacqua KG, Gottschlich A, Murchland AR, Alvarez CS, Rivera-Andrade A, Meza R. Cervical cancer knowledge and barriers and facilitators to screening among women in two rural communities in Guatemala: a qualitative study. BMC Women’s Health. 2022;22(1):197.

37. Lidofsky A, Miller A, Jorgensen J, Tajik A, Tendeu K, Pius D, et al. Development and implementation of a culturally appropriate education program to increase cervical cancer screening among Maasai women in rural Tanzania. Annals of Global Health. 2019;85(1):127.

38. Tillman S. Consent in pelvic care. Journal of Midwifery & Women’s Health. 2020;65(6):749–58.

39. Bateman LB, Blakemore S, Koneru A, Mtesigwa T, McCree R, Lisovicz NF, et al. Barriers and facilitators to cervical cancer screening, diagnosis, follow-up care and treatment: Perspectives of human immunodeficiency virus-positive women and health care practitioners in Tanzania. The Oncologist. 2019;24(1):69–75.

40. Shariati-Sarcheshme M, Mahdizdeh M, Tehrani H, Jamali J, Vahedian-Shahroodi M. Women’s perception of barriers and facilitators of cervical cancer Pap smear screening: a qualitative study. BMJ open. 2024;14(1):e072954.

